# Distinct age-specific SARS-CoV-2 IgG decay kinetics following natural infection

**DOI:** 10.1101/2021.08.05.21259465

**Authors:** Calvin P Sjaarda, Emily Moslinger, Kyla Tozer, Robert I. Colautti, Samira Kheitan, Robyn Meurant, Stefanie Van Cleaf, Ali Ardakani, Oliver Bosnjak, Abdi Ghaffari, Prameet M Sheth

## Abstract

**Background:** Antibody responses to SARS-CoV-2 can be observed as early as 14 days post-infection, but little is known about the stability of antibody levels over time. Here we evaluate the long-term stability of anti-SARS-CoV-2 IgG antibodies following infection with SARS-CoV-2 in 402 adult donors.

**Methods:** We performed a multi-center study carried out at Plasma Donor Centers in the city of Heidelberg (Plasmazentrum Heidelberg, Germany) and Munich (Plasmazentrum München, Germany). We present anti-S/N and anti-N IgG antibody levels in prospective serum samples collected up to 403 days post recovery from SARS-CoV-2 infected individuals.

**Results:** The cohort includes 402 adult donors (185 female, 217 male; 17 - 68 years of age) where anti-SARS-CoV-2 IgG levels were measured in plasma samples collected between 18- and 403-days post SARS-CoV-2 infection. A linear mixed effects model demonstrated IgG decay rates that decrease over time (χ^2^=176.8, p<0.00001) and an interaction of time*age χ (χ^2^=10.0, p<0.005)), with those over 60+ years showing the highest baseline IgG levels and the fastest rate of IgG decay. Baseline viral neutralization assays demonstrated that serum IgG levels correlated with *in vitro* neutralization capacity in 91% of our cohort.

**Conclusion:** Long-term antibody levels and age-specific antibody decay rates suggest the potential need for age-specific vaccine booster guidelines to ensure long term vaccine protection against SARS-CoV-2 infection.

## Main Text

The spread of Severe Acute Respiratory Syndrome Coronavirus-2 (SARS-CoV-2) now represents the largest outbreak of viral infectious disease in the last century. Since its discovery in late 2019(1), the virus has spread globally, shutting down industry, air travel and forcing governments to implement drastic public health measures, quarantines, curfews, and lockdowns in an effort to mitigate its spread(2). At the time of writing this article over 176 million confirmed cases of COVID-19 had been documented, and global vaccination programs were well-underway in an effort to control the virus.

Protection resulting from natural infection and vaccination programs all depend on eliciting a durable protective host immune response. The early antibody response to SARS-CoV-2 following infection is predominated by IgA, IgM and IgG responses that are all detectable as early as 7-14 days post infection(3, 4). Although robust CD4+ and CD8+ T cell immune responses are detectable in the first 30 days following infection(5), the best correlate of protection against SARS-CoV-2 remains the IgG immune response against the SARS-CoV-2 spike protein(6). Indeed, IgG adoptive transfer studies done in non-human primates (NHPs) found that purified IgG from infected NHPs protected naïve NHPs from infection(6). However, antibody responses to coronaviruses are highly variable. Studies in individuals infected with HCoV-OC43 and HCoV-229E demonstrate that antibody levels and protection from disease was short-lived with re-infection observed within 12 months(7, 8). On the other hand, antibody responses mounted against SARS-CoV appeared to be maintained at detectable levels for up to 2-years post infection(9). The long-term stability of antibodies against SARS-CoV-2 is not well understood, although early reports have demonstrated gradual decay in antibodies levels 6-9 months post infection(10). Understanding the dynamics of antibody levels following natural infections provides key insights into the potential for the development of long-term immunity against SARS-CoV-2 following both vaccination and natural infection.

We recruited 402 individuals with confirmed SARS-CoV-2 infection (PCR) and mild to moderate COVID-19 symptoms (Table 1). We tested an average of 9.8 blood samples per donor (range: 1-34) that were collected between 18 and 403 days after the suspected date of infection. The first blood sample collected following recovery was labeled as patient baseline. Over 92% (372/402) of individuals mounted detectable IgG immune responses against the SARS-CoV-2 spike (S) and nucleocapsid (N) proteins (measured by the YHLO iFlash IgG assay). The S-protein is the primary receptor binding site with putative mutations in the S-protein being associated with increased virulence and transmission efficiency(11, 12). In our cohort, biological sex was not associated with differences in baseline anti-spike protein IgG (Figure 1A); however, there was a significant difference in baseline antibody levels between age groups binned by decade (Kruskal–Wallis; p < 0.0001) (Figure 1B). Median baseline anti-spike/nucleocapsid IgG were lowest in individuals between 20-29 years, while individuals 60+ years had the highest antibody levels in sera.

**Table 1.**
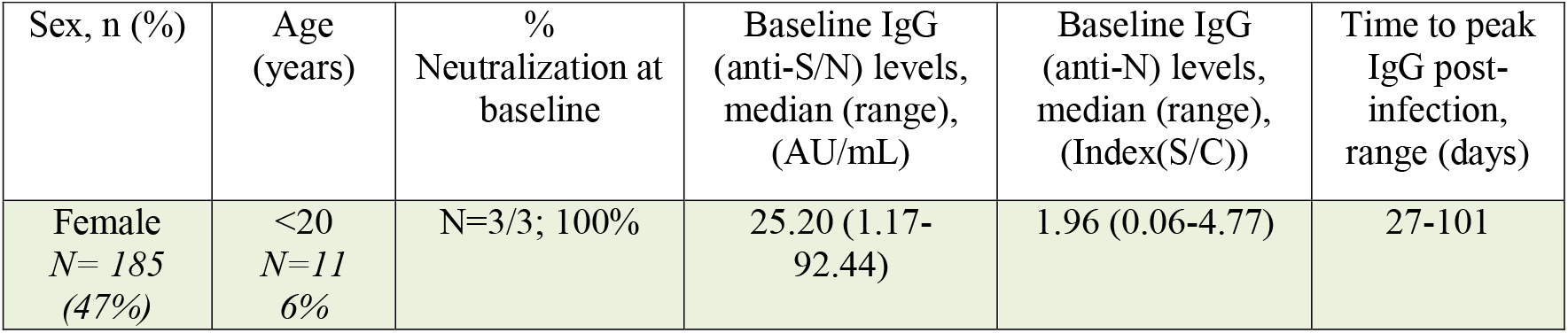
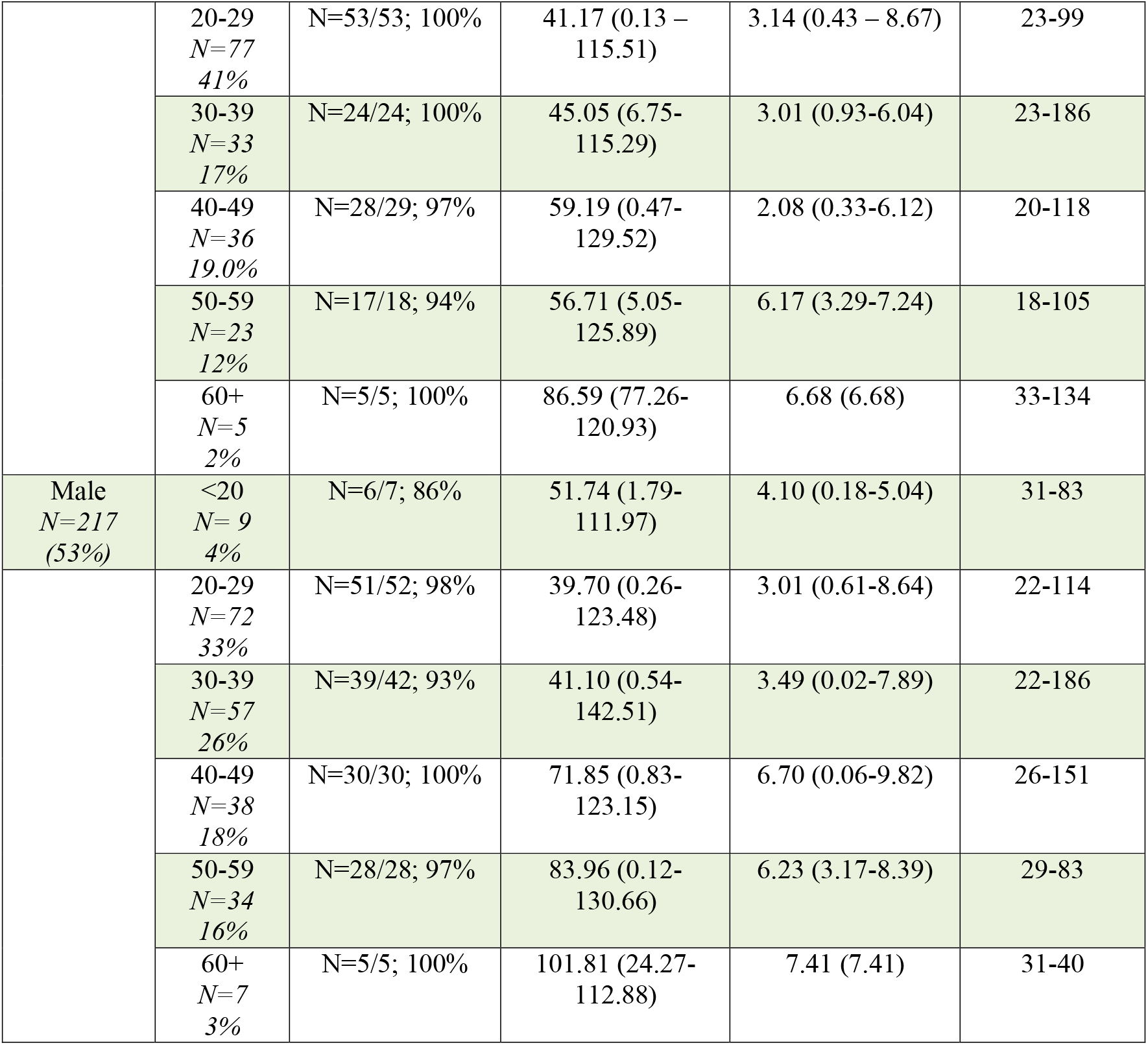
Characteristics of the study cohort

| Sex, n (%) | Age (years) | % Neutralization at baseline | Baseline IgG (anti-S/N) levels, median (range), (AU/mL) | Baseline IgG (anti-N) levels, median (range), (Index(S/C)) | Time to peak IgG post-infection, range (days) |
| --- | --- | --- | --- | --- | --- |
| Female<br>*N*= 185<br>(47%) | <20<br>*N*=11<br>6% | *N*=3/3; 100% | 25.20 (1.17-92.44) | 1.96 (0.06-4.77) | 27-101 |
|  | 20-29<br>N=77<br>41% | N=53/53; 100% | 41.17 (0.13 – 115.51) | 3.14 (0.43 – 8.67) | 23-99 |
|  | 30-39<br>N=33<br>17% | N=24/24; 100% | 45.05 (6.75- 115.29) | 3.01 (0.93-6.04) | 23-186 |
|  | 40-49<br>N=36<br>19.0% | N=28/29; 97% | 59.19 (0.47- 129.52) | 2.08 (0.33-6.12) | 20-118 |
|  | 50-59<br>N=23<br>12% | N=17/18; 94% | 56.71 (5.05- 125.89) | 6.17 (3.29-7.24) | 18-105 |
|  | 60+<br>N=5<br>2% | N=5/5; 100% | 86.59 (77.26- 120.93) | 6.68 (6.68) | 33-134 |
| Male<br>N=217<br>(53%) | <20<br>N= 9<br>4% | N=6/7; 86% | 51.74 (1.79- 111.97) | 4.10 (0.18-5.04) | 31-83 |
|  | 20-29<br>N=72<br>33% | N=51/52; 98% | 39.70 (0.26- 123.48) | 3.01 (0.61-8.64) | 22-114 |
|  | 30-39<br>N=57<br>26% | N=39/42; 93% | 41.10 (0.54- 142.51) | 3.49 (0.02-7.89) | 22-186 |
|  | 40-49<br>N=38<br>18% | N=30/30; 100% | 71.85 (0.83- 123.15) | 6.70 (0.06-9.82) | 26-151 |
|  | 50-59<br>N=34<br>16% | N=28/28; 97% | 83.96 (0.12- 130.66) | 6.23 (3.17-8.39) | 29-83 |
|  | 60+<br>N=7<br>3% | N=5/5; 100% | 101.81 (24.27- 112.88) | 7.41 (7.41) | 31-40 |

**Figure 1.**
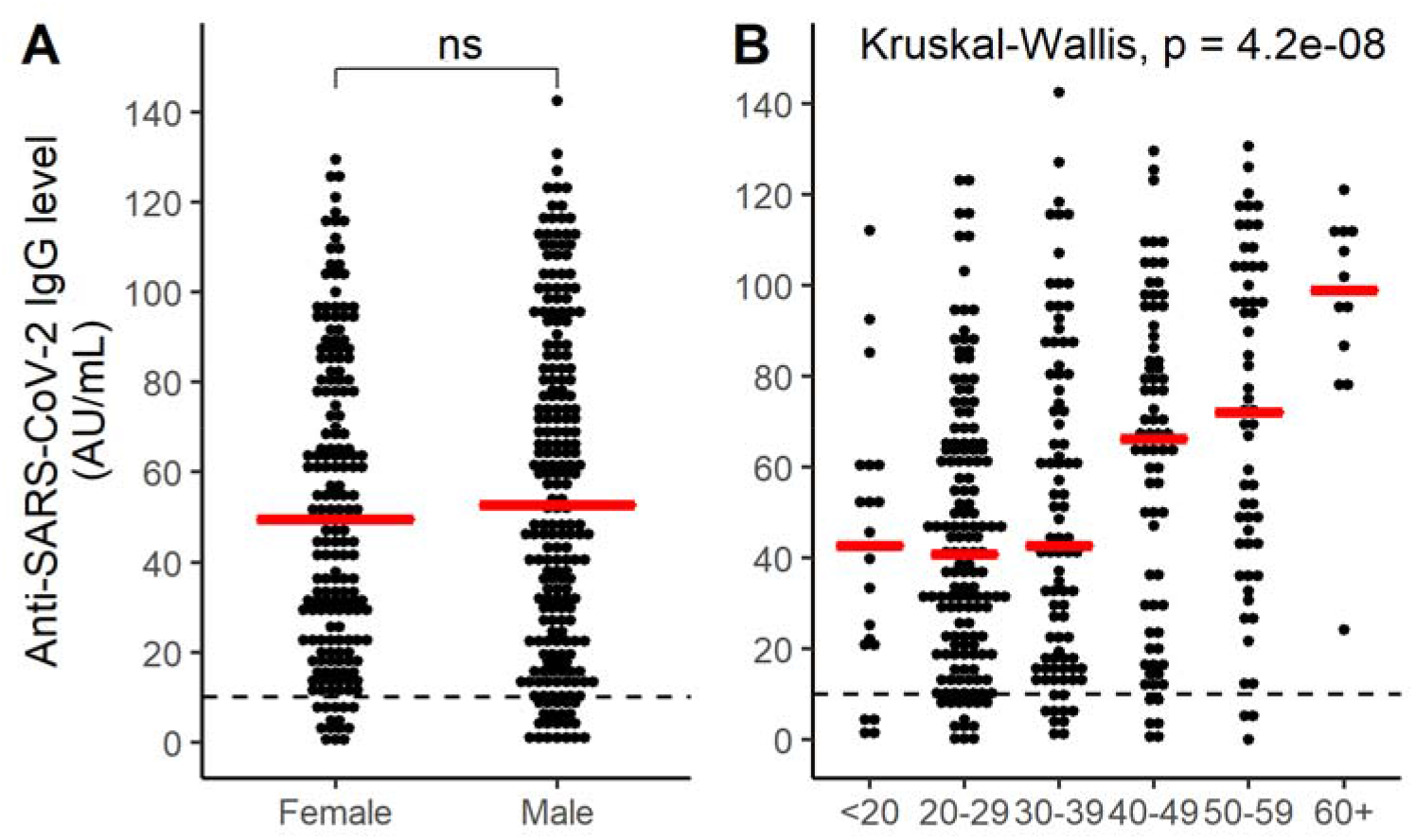
Baseline serum anti-S/anti-N IgG levels. There was no significant difference in baseline median IgG levels, measured by iFlash test, in males and females (A). Age stratification of the study cohort identified a statistically significant difference with baseline IgG levels between age groups (B).

To support our IgG measurements, we validated the data using a second IgG test (Abbott Architect, anti-N IgG). We performed a linear mixed effects model to determine decay kinetics of anti-S/anti-N (iFlash) and anti-N (Architect) IgG antibodies. Donors with less than 3-prospective blood samples were removed resulting in a final dataset of 296 donors and 2392 individual IgG measurements for the iFlash assay and 162 donors and 1084 measurements for the Architect assay. To account for random effects of donor, we normalized individual responses using loge IgG – mean (loge IgG) and log-transformed the time since infection with SARS-CoV-2 (Fig S1 and Fig S1). Details of the model and analysis are presented in the Supplementary Materials.

A linear mixed effect model demonstrated that IgG levels decline significantly over time (Fig. S3; χ^2^ = 176.8, p <0.0001), with decay rates that vary significantly with the age of the participant χ^2^= 10.0, p <0.005). To visualize the interaction between time and age, we binned participant age by decade (Figure 2). Individuals in the highest age class (60+) had the fastest rates of IgG decay (slope; iFlash = -0.59 and Architect = -0.79), while individuals in the age groups 40 – 49 and 50 – 59 had the slowest average rate of decay in both assays compared to all other age groups (Figure 2). We did not detect an effect of biological sex on IgG levels or decay rate in our cohort (Fig. S4). Although the assays have different targets (anti-S/anti-N vs. anti-N IgG), antibodies to both regions of the virus had comparable decay kinetics with IgG levels falling to below the limit of assay detection after 125.5 days (95% CI 116.8-134.4) for the iFlash assay and 132.2 days (95% CI 119.6-144.9) for the Architect assay (Fig. S5).

**Figure 2.**
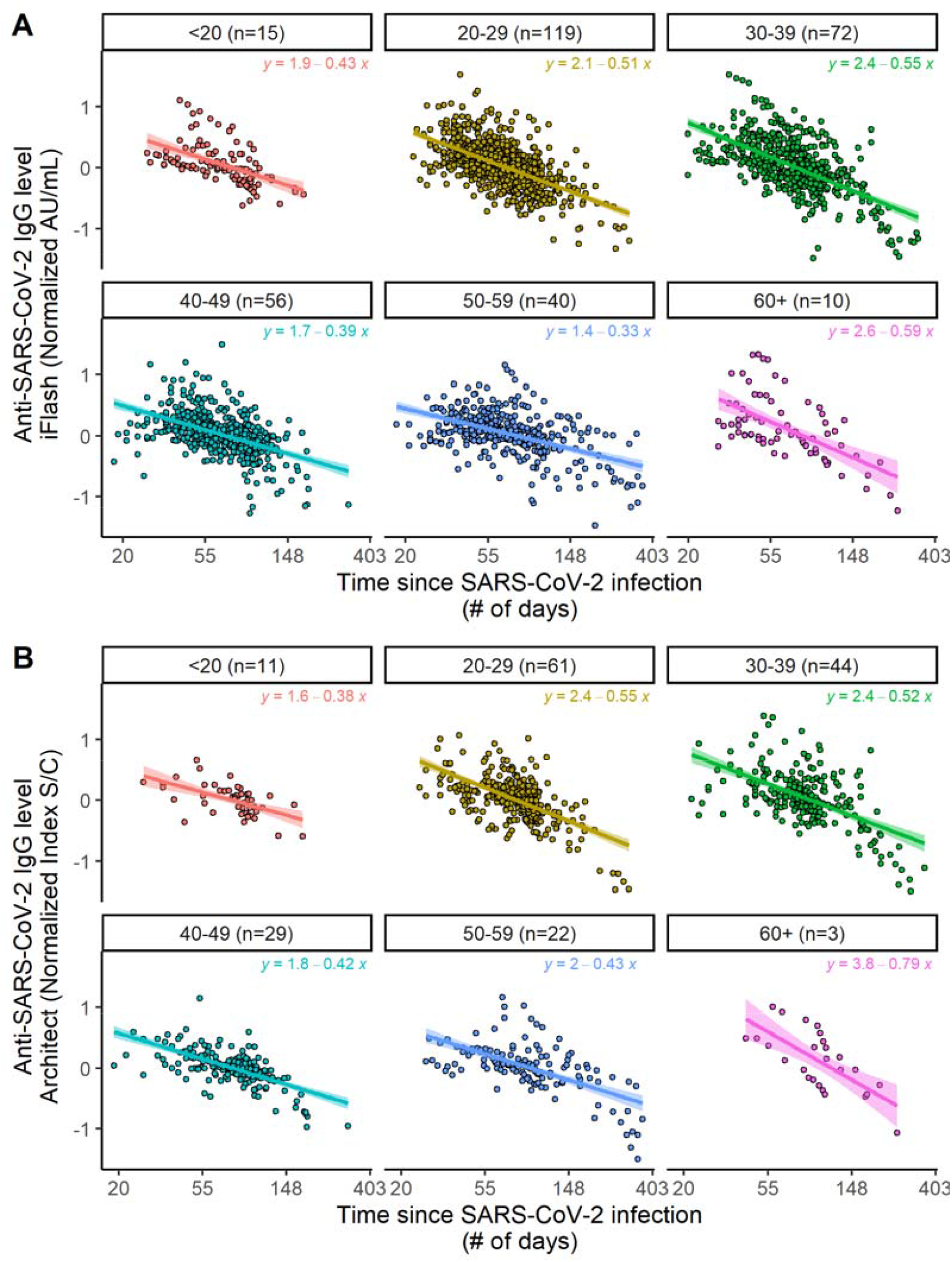
Age stratified IgG decay kinetics. IgG antibody levels, measured by iFlash (anti-S/anti-N) (A) and Architect (anti-N) (B) assays, demonstrate significant anti-SARS-CoV-2 IgG antibody decay over time that is dependent on the age of the participant. The equations for lines of best fit and the number of subjects per age groups are indicated at the top of each plot. Variability between donor was normalized on the y-axis by taking the loge IgG – mean (loge IgG) for each participant.

IgG titers in the sera of SARS-CoV-2 infected patients (up to 94 days post onset of symptoms) and those receiving vaccinations (within 3 weeks after first dose) have been used as surrogate markers of immunity. Similarly, vaccine studies often cite IgG titers against viral spike protein as a measure of vaccine protection (13-15) including in response to emerging variants of concern(16). However, linking IgG titers to viral immunity requires an understanding of the absolute titers of anti-spike IgG that are required for protection. To evaluate the relationship between serum anti-S/anti-N IgG levels and the capacity of patient sera to neutralize SARS-CoV-2 infection in vitro, baseline neutralization assays were performed in 279/402 patients within 100 days of testing positive for SARS-CoV-2. Neutralization capacity was observed in the sera of 273/279 (97.8%) of patients. Serum of four patients (1.3%) did not neutralize SARS-CoV-2 in vitro, despite detectable levels of anti-S/N IgG. Eighteen patients (6.1%) neutralized SARS-CoV-2 in vitro despite having no detectable anti-S/N IgG in sera (Figure 3). These data suggest that in our cohort the anti-S/anti-N IgG levels (measured by iFlash test) is a good surrogate marker of protective immunity against SARS-CoV-2 infection in most, but not all individuals.

**Figure 3.**
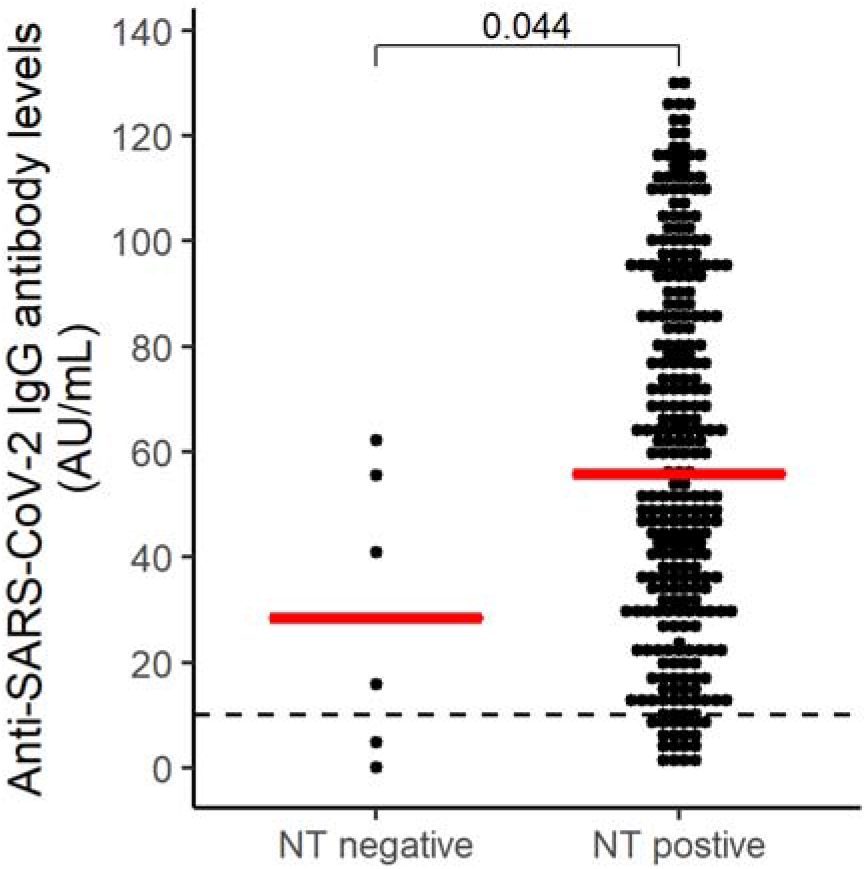
Serum IgG levels as a surrogate marker of protective immunity as determined by neutralization assay. The results of virus neutralization assay, performed at baseline, (NT) is plotted against anti-S/anti-N IgG levels (iFLash test). The p value represents the difference between median IgG levels between NT positive and negative cases at baseline (two-tailed non-parametric Mann-Whitney U test). Dashed line represents the limit of detection (LOD) of iFlash IgG test.

Using long-term (>400d) sequential serum samples from recovered COVID-19 patients, our results demonstrate a rapid decay of IgG levels in all age groups with the fastest decay rates observed in 60+ year-old adults. Collier et al. also reported heterogeneity in immune response participants of different ages following vaccination where older participants had lower antibody levels (quantity) and/or lower antibody affinity (quality) (17). This study does not evaluate the potential role in viral immunity played by non-IgG immune responses including innate and adaptive cell mediated responses that have been implicated in protection (6). Although antibody responses against SARS-CoV-2 have been seen to be longer lived following mRNA vaccination compared to natural infection (14), the rapid decay in IgG observed in our study suggests that further longitudinal studies are needed to accurately predict long-term immunity in vaccinated and infected individuals. If anti-S/anti-N IgG titers are a good surrogate for protective immunity against SARS-CoV-2, then vaccine boosters will be required by the second year of post-infection recovery and/or immunization to protect against re-infection. Further, our data suggests the optimal timeline for vaccine booster may depend on the age of the patient, for example by prioritizing groups with more rapid decay of antibodies.

## Data Availability

All data are available in the main text or the supplementary materials. The data analysis pipeline is available at https://github.com/CalvinSjaarda/COVID_immune_IgG.git.

## Author contributions

Conceptualization: OB, RM, AA, AG, PMS, Methodology: OB, CPS, Investigation: OB, RM, AA, Visualization: CPS, RC, PMS, AG, Funding acquisition: OB, AA, Project administration: AG, PMS, Supervision: AG, PMS, OB, Writing – original draft: CPS, EM, KT, Writing – review & editing: CPS, EM, KT, PMS, AG

## Ethics

All donors provided informed consent and had to meet the German Medical Association criteria for blood and blood component collection according to the transfusion and hemotherapy guidelines. All experimental protocols were approved by and conducted in accordance with the Ethics Commission of the Bavarian State Medical Association (No 05142).

