## Supplementary Materials for "Distinct age-specific SARS-CoV-2 IgG decay kinetics following natural infection"

Materials and Methods

**Study Design**

We performed a multi-center study carried out at Plasma Donor Centers in the city of Heidelberg (Plasmazentrum Heidelberg, Germany) and Munich (Plasmazentrum München, Germany). The target population for the study were SARS-CoV-2 recovered adults that were chosen for plasma donation by plasmapheresis. All donors provided informed consent and had to meet the German Medical Association criteria for blood and blood component collection according to the transfusion and hemotherapy guidelines(1). Samples were collected in a longitudinal manner and processed in the Biomex GmbH Laboratory and Laboratory Limbach for serological analysis (Heidelberg, Germany).

**Patient recruitment and sample preparation**

Between March 30, 2020 and April 9, 2021, plasma samples were collected from 402 adult donors (185 female, 217 male) that recovered from COVID-19 and referred to Plasma Collection Centers in Heidelberg and Munich. Documented donor information included age, biological sex, country of birth, time and place of suspected infection, date of positive SARS-CoV-2 PCR test, date of quarantine, and clinical symptoms. The SARS-CoV-2 infections were confirmed by nasopharyngeal swab and PCR based on the test guidelines of the German National Laboratory for Coronaviruses (Charité–Universitätsmedizin Berlin, Germany). Plasma samples were collected by plasmapheresis using sodium citrate 4% w/v anticoagulant solution at room temperature. Samples were taken from the plasma bottle using vacuettes without additives. These were frozen with the plasma bottle at < -20°C for subsequent serological analysis.

**Serological Analysis**

The measurements of IgG against SARS-CoV-2 in plasma samples were performed using iFlash 1800 paramagnetic particle chemiluminescent immunoassay (CLIA) from Shenzhen YHLO Biotech (Shenzhen, China) and Architect chemiluminescent microparticle immunoassay (CMIA) from Abbott Laboratories (IL, USA). The iFlash assay detects antibodies against SARS-CoV-2 spike (S) and nucleocapsid (N) proteins and the Architect assay detected antibodies against the N protein. All plasma antibody tests were performed with fully automated analyzers according to the manufacturer’s instructions. Antibody levels were calculated and results interpreted according to the manufacturer’s instructions and defined cut-offs for antibody positivity.

***Neutralization assay***

Samples were screened for neutralizing antibodies using the SARS-CoV-2 sVNT ELISA (GenScript, New Jersey, USA) according to the manufacturer’s instructions. Samples below 30% inhibition were confirmed using the WANTAI SARS-CoV-2 Ab ELISA (Beijing Wantai Biological Pharmacy Enterprise; Beijing, China) for detection of complete antibodies against SARS-CoV-2 according to manufacturer’s instructions, with the exception that we applied 50 µl of sample. Additionally, sVNT ELISA negative samples (below 30% inhibition) were confirmed in a biological neutralisation test (NT) as follows: a total of 100µL of plasma sample was diluted in DMEM (10 % FCS, 2 mM L-glutamine) in six 2-fold dilutions resulting in dilutions of 1:10 up to 1:320. Dilutions were mixed 1:1 with SARS-CoV-2 (strain BetaCoV/Germany/BavPat1/2020, kindly provided by Dr. Roman Woelfel, Institute for Microbiology of the German Armed Forces; final virus concentration 1,000 TCID_50_ /mL) and incubated at room temperature for 1 h. Subsequently, 100 µL of diluted serum-virus mix were added to wells containing 2 x 10^4^ Vero E6 cells per well (#85020206, European Collection of Authenticated Cell Cultures, Porton Down, UK), in a 96-well plate. Each sample dilution was tested in eight replicates and cells were incubated for 5 days at 37 °C, 5% CO_2_. After 5 days, each well was analysed by light microscopy for visible cytopathic effect (CPE). The number of wells without CPE (negative wells) was counted and PRNT_50_ values were calculated as previously described. For quality control, a positive control with defined titer was analysed in parallel and back-titration of the virus stock was performed. Samples were considered positive for neutralising antibodies when NT (titer ≥ 1:15) or sVNT ELISA (> 31% inhibition) was positive.

**Statistics**

We used the lme4 (v1.1-26) package(2) in R version 4.0.3 (3) to fit the data to a linear mixed effect (lme) model. We fitted sequential models to include main effects of time since infection (log transformed, continuous variable), age (continuous variable), biological sex (factor), and assay (iFlash or Architect as a factor) and interactions between main effects as fixed effects and the intercepts and slopes for the donor as random effects. The model that best fit the data (determined by anova) included significant main effect of time (χ^2^=176.8, p<0.00001), assay (χ^2^=344.42, p<0.00001), significant interactions of time*age (χ^2^=10.0, p<0.005) and time*assay (χ^2^=5.27, p<0.05), and significant random effect of donor intercept (χ^2^=117.48, p<0.00001) and donor slope (χ^2^=189.27, p<0.00001).

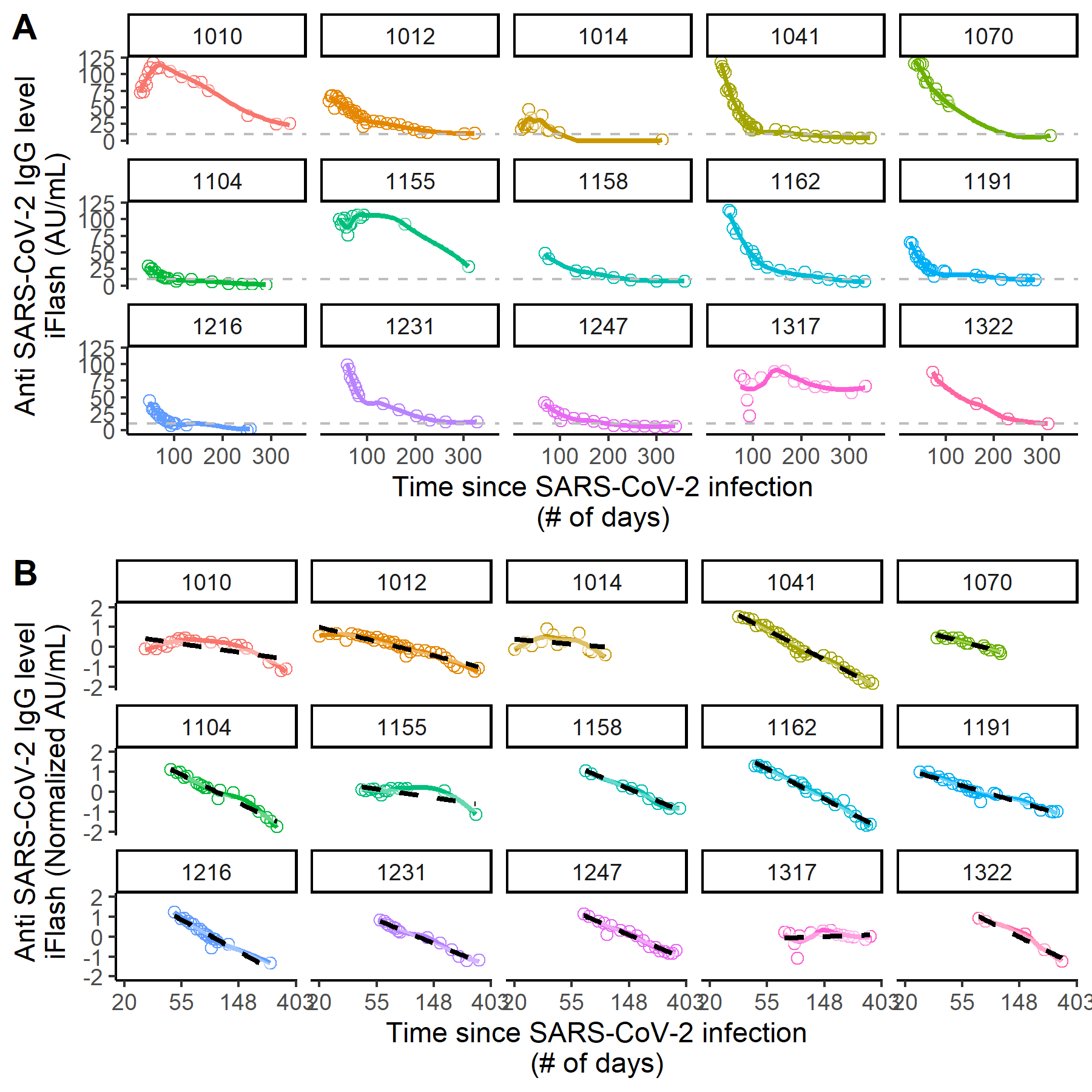

Fig. S1. Individual IgG antibody levels determined using the iFlash assay for fifteen participants with repeated sample collection dates (mean = 17.73 samples per donor). A. Individuals demonstrate considerable variation in their scale of production and decay rates of IgG antibodies for SARS-CoV-2 (detection limit of assay is shown by dashed grey line). B. Normalization and log transformation of IgG levels for each participant log transformation of time since infection enables a predicted decay rate provided by the linear mixed effects model (shown by dashed black line).

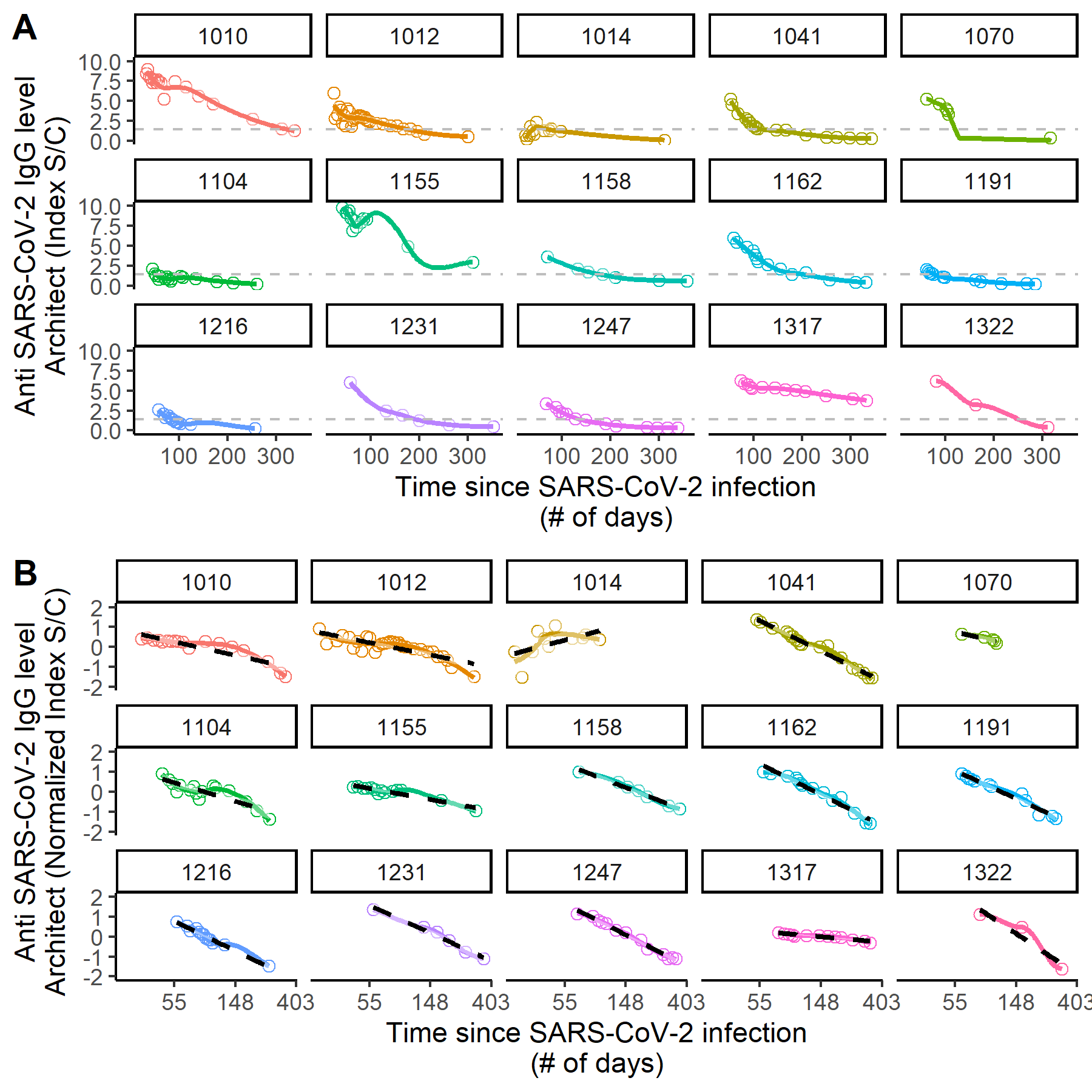

Fig. S2. Individual IgG antibody levels determined using the Architect assay for fifteen participants with repeated sample collection dates (mean = 13.13 samples per donor). A. Individuals demonstrate considerable variation in their scale of production and decay rates of IgG antibodies for SARS-CoV-2 (detection limit of assay is shown by dashed grey line). B. Normalization and log transformation of IgG levels for each participant and log transformation of time since infection enables a predicted decay rate provided by the linear mixed effects model (shown by dashed black line).

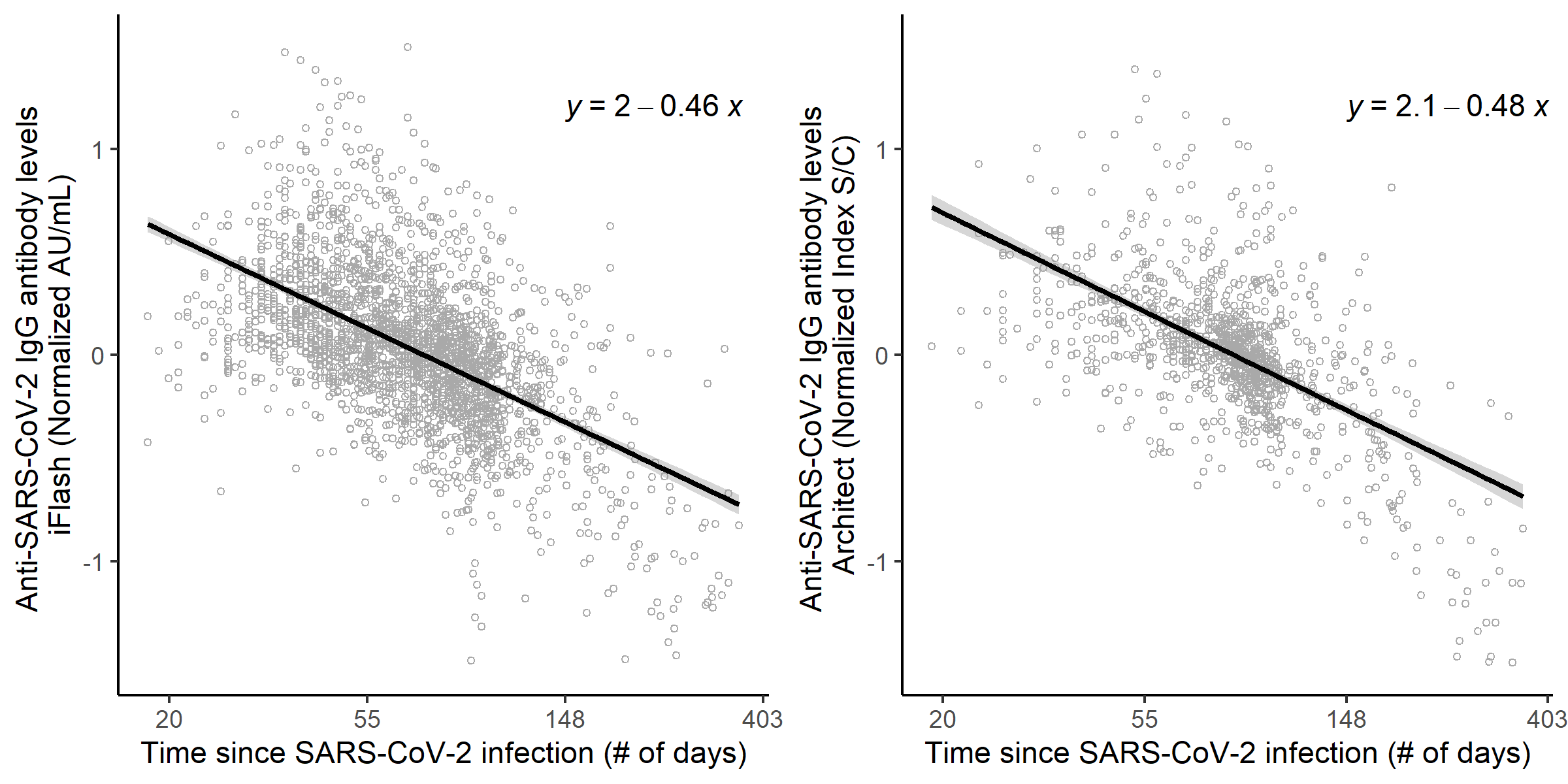

Fig. S3. IgG decay rates over 400 days. iFlash (YHLO, n=312) and Architect (Abbot, n=170) IgG assays demonstrated that anti-SARS-CoV-2 IgG antibodies decayed over time at similar rates with almost identical decay slopes (-0.46 vs. -0.48). Variability between donor was normalized on the y-axis by taking the log_e_ IgG – mean (log_e_ IgG) for each participant.

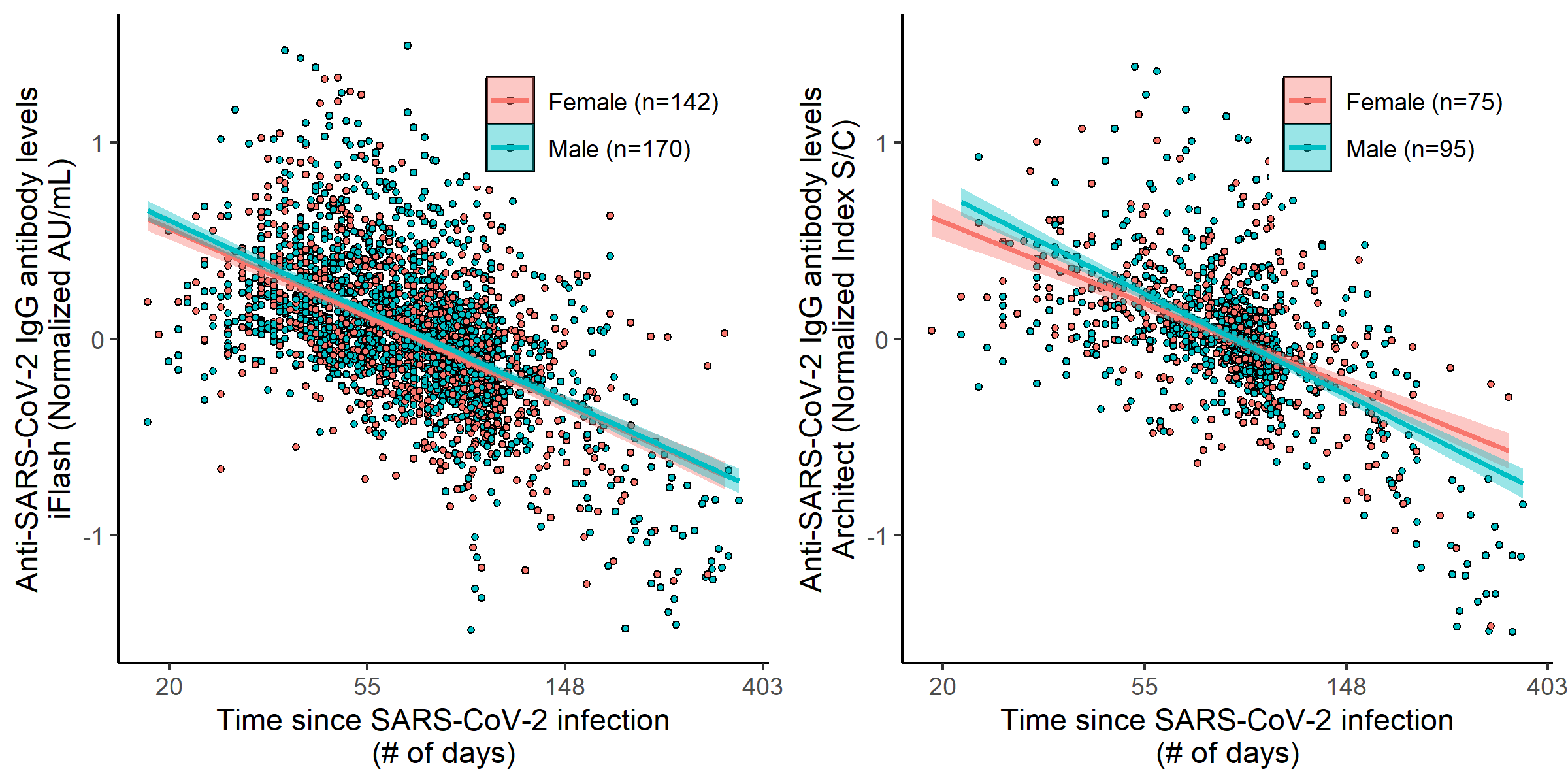

Fig. S4. Biological sex did not have a significant effect on detection level or the decay rate of IgG in infected individuals over time (χ^2^=0.053, p=0.8178) seen in both the Architect (A) and iFlash (B) assays. Variability between donor was normalized on the y-axis by taking the log_e_ IgG – mean (log_e_ IgG) for each participant.

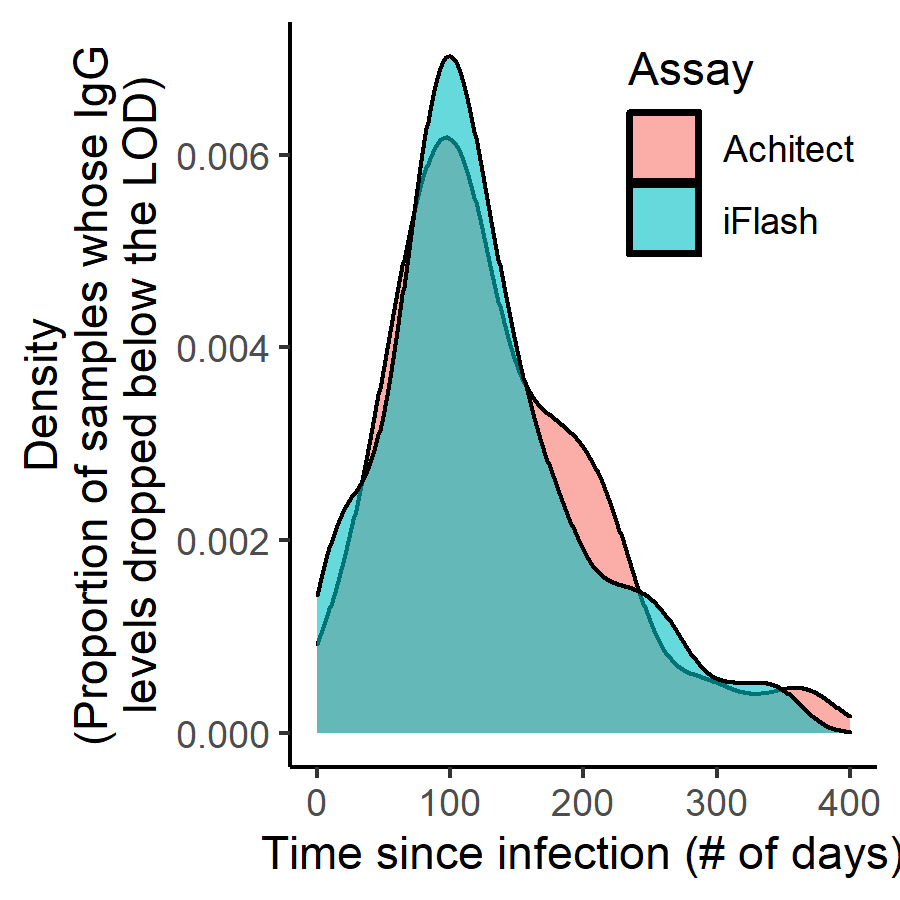

Fig. S5. Distribution of the time predicted for donor IgG levels to decay below the limit of detection.

3.	R Core Team. R: A language and environment for statistical computing. Vienna, Austria: R Foundation for Statistical Computing; 2020.
